# Neural Alterations in Chronic Pain: MRI Analysis

**DOI:** 10.64898/2026.08.05.26359702

**Authors:** Lital Cohen-Blum, Shaked Eizman, Pascal Tétreault, Or Duek

## Abstract

**Background:** Chronic pain affects hundreds of millions worldwide and remains a major clinical challenge, despite numerous available treatments. Advances in brain imaging offer a promising path toward identifying neural signatures of chronic pain, potentially enhancing diagnosis and guiding treatment. However, while a core set of brain regions, including the insula, cingulate, and somatosensory cortices, has been repeatedly implicated, findings regarding other regions and connectivity patterns involved remain inconsistent, with limited robust replication.

**Objective:** To address these gaps, the present work characterizes resting-state functional connectivity and gray matter volume differences between chronic pain patients and pain-free controls.

**Methods:** In this secondary analysis of publicly available data, anatomical and resting-state functional MRI were analyzed from 56 patients with chronic knee pain due to osteoarthritis and 20 pain-free controls. Group comparisons used Network-Based Statistic (NBS) and Bayesian multivariate regression models, controlling for demographic covariates.

**Results:** In the pain group, about 75% of parcellated brain regions exhibited increased functional connectivity compared to controls. The 30 highest degree centrality regions in the NBS network were concentrated in regions consistent with prior pain neuroimaging findings. Additionally, chronic pain patients exhibited reduced gray matter volume (-3.98%; SD 1.2%) across 33% of parcellated brain regions, including key regions implicated in pain processing.

**Conclusions:** These findings demonstrate widespread functional and anatomical neural alterations in chronic pain, revealing a global pattern of reorganization extending beyond previously reported network-pair effects. Characterizing such alterations may contribute to ongoing efforts to identify neuroimaging markers of chronic pain, with potential translational relevance.

## INTRODUCTION

Chronic pain poses a significant global challenge, leaving millions debilitated and unanswered. Typically defined as pain persisting longer than three months ^1^, it manifests in physical and psychological symptoms, including reduced mobility, anxiety, and disruptions in sleep and appetite ^2^, all of which significantly compromise quality of life. Chronic pain affects 20%-40% of adults across different populations ^3–7^, making it the most common reason for seeking medical attention ^8^. Yet despite its prevalence, current treatments are suboptimal, highlighting the need for improved objective biomarkers and a mechanistic understanding of chronic pain.

### Neural mechanisms

Chronic pain is currently diagnosed primarily through subjective reports of its intensity and location, as no FDA-approved biomarkers exist. This lack of objective, quantitative tools for diagnosing, tracking progression, or evaluating treatment responses limits effective medical decision-making ^9^. Neuroimaging offers a promising avenue for identifying objective markers of chronic pain, though translation to clinical practice remains a challenge. Analyzing Magnetic Resonance Imaging (MRI) datasets from chronic pain patients offers a promising path forward, yielding insights into the brain’s functional connectivity and anatomical disruptions that could ultimately enable more effective, personalized treatment.

Pain is a subjective experience shaped by complex sensory, emotional, and cognitive processes ^10^, and is increasingly investigated using MRI. Early models, like the “pain neuromatrix” emphasized limbic and thalamocortical involvement ^11,12^, while newer frameworks, such as the “dynamic pain connectome” ^13^ describe pain as emerging from shifting interactions across brain networks. This framework emphasizes the Default Mode Network (DMN), salience network, and ascending nociceptive and descending pain modulation systems ^14^. The transition from acute (short-term) to chronic pain involves substantial neural reorganization ^15–17^. Gray matter reductions extend beyond sensory-motor regions to areas supporting interoception, beliefs, rewards and motivation, such as the posterior cingulate, Ventromedial Prefrontal Cortex (vmPFC), nucleus accumbens and insula ^15,18^. This aligns with evidence that the acute-to-chronic transition reflects a shift away from nociception, toward higher-order integrative processes ^18–21^. Furthermore, structural and functional connectivity alterations in pain-related regions have been repeatedly observed ^15,16^, with key regions including those above, plus the Anterior Cingulate Cortex (ACC), and thalamus ^17^. Despite these advances, the precise neural pathways underlying chronic pain remain incompletely understood, highlighting a critical knowledge gap.

Resting-state functional MRI (rs-fMRI) is a widely used technique that measures spontaneous fluctuations in Blood Oxygen-Level Dependent (BOLD) signals across the brain, without an explicit task ^22^. It enables the identification of resting-state networks, brain regions exhibiting correlated activity at rest. Disruptions in resting-state functional connectivity have been reported across neurological and psychiatric disorders, and the “connectome”, representing the covariance structure of brain activity, is increasingly regarded as a promising candidate disease biomarker ^22–24^. While methodological limitations have been noted, with some authors calling for more interpretable designs ^25,26^; rs-fMRI remains a non-invasive and promising tool for identifying neural markers of chronic pain ^27^. Building on prior evidence of neural alterations in chronic pain, we use rs-fMRI and structural MRI to characterize whole-brain functional connectivity and gray matter volume differences between chronic pain patients and pain-free controls.

## METHODS

### Participants

The current study is a secondary analysis from previous studies ^28,29^ using an open dataset available at https://openneuro.org/datasets/ds000208/versions/1.0.1 or at https://openpain.org/ (dataset named PlaceboPrediction). Out of 143 chronic pain patients with knee osteoarthritis recruited across 3 studies, 56 completed brain scans in studies 1 and 2. Additionally, 20 age-matched pain-free control participants were recruited to match with 17 patients from study 1. Since the groups had similar mean ages and comparable sex distribution, the two samples were merged into a single pain group (N=17+39=56), and compared with pain-free controls (N=20). Inclusion criteria for chronic pain patients: pain persisting for at least 1 year and a pain intensity rating of 4/10 or higher within 48 hours prior to the screening visit (refer to supplement A for demographic and pain scores table).

#### Ethics Statement

This study is a secondary analysis of previously collected data from studies approved by the Northwestern University Institutional Review Board (STU00039556 for studies 1 and 2; STU00059872 for study 3) and registered on ClinicalTrials.gov (NCT02903238 and NCT01558700). All participants provided written informed consent prior to participation in the original studies.

### Tools

#### MRI acquisition

Anatomical T1-weighted images (MP-RAGE) were acquired with a 3T Siemens Trio scanner (1 mm^3^ voxels, TR=2500ms, TE=3.36ms, 160 slices). T2-weighted resting-state fMRI data were collected on the same scanner (TR=2500 ms, echo time TE=30 ms, 40 slices, 3 mm thickness, 300 volumes) covering the whole brain.

#### Questionnaires

Patients participating in studies 1 and 2 filled out a general health questionnaire and a Visual Analog Scale (VAS) for their knee pain, rated from 0 to 10. Additionally, they completed the Western Ontario and McMaster Universities Osteoarthritis Index (WOMAC), the Beck Depression Inventory (BDI), and the Pain Catastrophizing Scale (PCS) questionnaires. All questionnaires were administered on the day of the brain scan.

### Design and results: *original* study

Design: The research comprised three studies: 1. a single-blind placebo pill trial (2 weeks), 2. a double-blind randomized placebo vs. duloxetine trial (3 months), and 3. an observational cohort without treatment or scans. Participants in studies 1 and 2 were fMRI-scanned *before treatment only*. Data from study 1 were used to identify predictors of placebo response, which were then validated in study 2. Results: Increased connectivity of the right mid-frontal gyrus emerged as the strongest predictor of placebo response in study 1 and accurately identified placebo responders in study 2 (95% accuracy) ^28^.

### Data analysis: *current* analysis

#### fMRI Preprocessing

##### fMRIPrep ^30^

A robust Python-based pipeline for fMRI preprocessing, including time shift correction, regression of head motion and other nuisance regressors, spatial smoothing, and frequency filtering ^31^. fMRIPrep version-24.0.1, default parameters were applied (refer to supplement D for details).

##### Parcellation

The Dictionary of Functional Modes for brain imaging (DiFuMo atlas ^32^, 256 regions) was used to extract functional regions of interest. Data from the cerebellum were excluded due to high levels of noise and uncertainty regarding its reliability, as reported by the study’s conductor. In addition, CSF regions were removed from the atlas, resulting in 219 brain regions.

##### Noise Regression

A general linear model (GLM) was applied to regress out noisy timepoints. Thresholds were set at Framewise Displacement (FD) average>0.5 and Derivative of Variance (DVARS)>3. Eight participants from the pain group were removed from functional analysis due to excessive noise (averaged FD>0.6) or missing data. No participants were excluded from the control group, resulting in a final sample of 48 participants in the pain group and 20 in the control group.

#### Functional analysis

Functional connectivity matrices ^33^ were extracted from the rs-fMRI data using the Python library ‘Nilearn’, version 0.10.1. Following time-series extraction and signal cleaning (as reported above), all regions’ time-series pairs were correlated using Pearson’s r correlation, resulting in 219×219 correlation matrices for each participant. Correlation matrices were Fisher-z transformed ^34^, to stabilize the variance of correlation coefficients.

#### Network-based statistic (NBS) and permutation testing ^35^

NBS is an analysis method that detects pairwise associations that differ substantially between groups. It incorporates permutation testing and can increase statistical power. The process is executed as follows: 1. Connectivity matrices are generated for each participant. 2. A t-test statistic is performed between the two groups. 3. The t-test is then thresholded to generate a set of suprathreshold (>threshold) links, creating a binary matrix named ‘adjacency matrix’. The network size, defined by the number of links, is recorded. Permutation testing is conducted by randomly shuffling pain group labels across participants while keeping the connectivity matrices fixed. For each of the M permutations, the group labels are reassigned, the test statistic is recalculated, and the same threshold is applied to identify suprathreshold links. The network size obtained for each permutation is recorded, and the observed network size is evaluated relative to the permutation-derived values to obtain a family-wise error-controlled *p*-value. The p-value for an observed network size is estimated by counting B, the number of permutations where the network size exceeds the observed one, then normalizing by M, the number of permutations (M=5000). Multiple t-value thresholds were tested for robustness: 2.5, 3, 3.5, and 4.

Following NBS, the degree centrality was calculated to characterize the connectivity structure of the identified network. Degree centrality values are proportional to the number of connections per node. Results focus on the 30 regions with the highest centrality values, highlighting nodes that act as major hubs within the network identified by NBS.

#### Anatomical analysis: gray matter volume

Gray matter volume was calculated using FreeSurfer ^36,37^, version 7.3.2. Freesurfer is a set of widely used and validated tools designed for neuroimaging analysis ^38–40^. The recon-all command of FreeSurfer, used for brain extraction, also includes the segmentation of white and gray matter ^41^. To compare chronic pain patients vs controls, a whole-brain Bayesian multilevel regression model was used on all 131 regions of FreeSurfer’s atlas, incorporating group differences and known confounding variables of age, sex, and total intracranial volume.

#### Bayesian analysis

Traditional statistical methods have faced increasing scrutiny due to replication challenges and limitations in interpreting p-values under null hypothesis significance testing. Bayesian inference offers an alternative framework with more direct interpretability and explicit incorporation of prior assumptions ^42^. Recent advances in computational power now enable efficient and robust Bayesian analysis of complex data. Bayesian inference quantifies uncertainty in model parameters by treating their values as probability distributions. Partially informative priors ^43^ were specified to encode plausible parameter ranges and were updated with the observed data to yield *posterior distributions*, which represent the probability of different parameter values given the data. The posterior assigns higher probability to values that better explain the data, and its spread reflects the remaining uncertainty; with limited data, posterior distributions are broad, becoming more concentrated as evidence increases ^44^. For continuous parameters, certainty can be summarized using Highest Density Interval (HDI) ^45^; A 94% HDI, the ArviZ default ^46^, was used, a deliberately non-conventional value chosen to emphasize the arbitrariness of any fixed cutoff ^47^. Results were considered robust when the HDI lay entirely above or below zero ^43^.

#### Additional analyses

Additional analyses are described in Supplementary Material C.

## RESULTS

### Functional analysis

Network-based statistic (NBS) analysis revealed an identified functional network showing stronger positive correlations across multiple brain regions in the pain group compared to pain-free controls. The pattern and strength of these correlations between brain regions are visualized in Figure 1A using a glass brain representation. Figure 1B presents the mean absolute correlation strength across all regions of the identified network, presented by both the individual participant level and the averaged pain group level.

**Figure 1:**
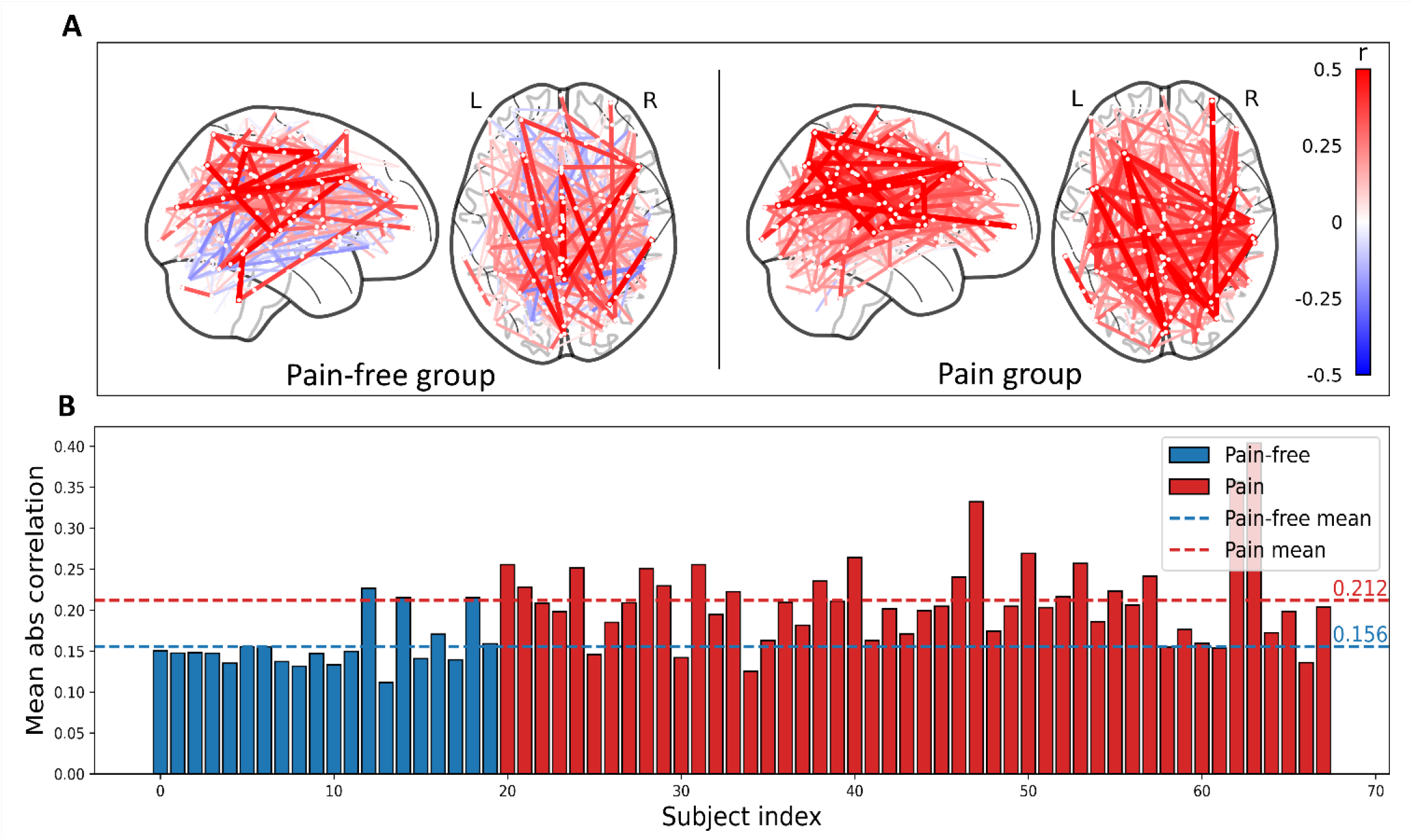
Increased functional connectivity in the chronic pain group. **(A) Pain group differences-significant edges according to NBS** Pairwise correlations between brain regions for the chronic pain group (right) and pain-free controls (left). Warmer colors indicate stronger positive correlations. Compared with controls, the pain group shows globally stronger connectivity. Network-based statistics identified a large subnetwork comprising **75% of brain regions** (166 of 219 nodes) with significantly stronger correlations in the pain group (t ≥ 3). Permutation test indicated network size significance (p = 0.009). **(B) Mean absolute correlation in identified NBS network** Distribution of mean absolute correlation values for all region pairs across participants. The chronic pain group shows higher overall connectivity (red; **mean** |**r**| **= 0.212**) compared with pain-free controls (blue; **mean** |**r**| **= 0.156**), consistent with the network-level differences shown in (A).

Figure 2 displays the 30 highest degree centrality values. These values are proportional to the number of connections each node (brain region) has with other nodes. Many of the highest degree centrality nodes of the found network corresponded to regions previously implicated in pain processing literature. According to Yeo’s 7-network parcellation, 10 of the highest 30 regions belong to the somatomotor network, while the remaining regions are distributed across the default mode network (DMN), salience, and dorsal attention networks (based on network labels derived from the DiFuMo atlas).

**Figure 2:**
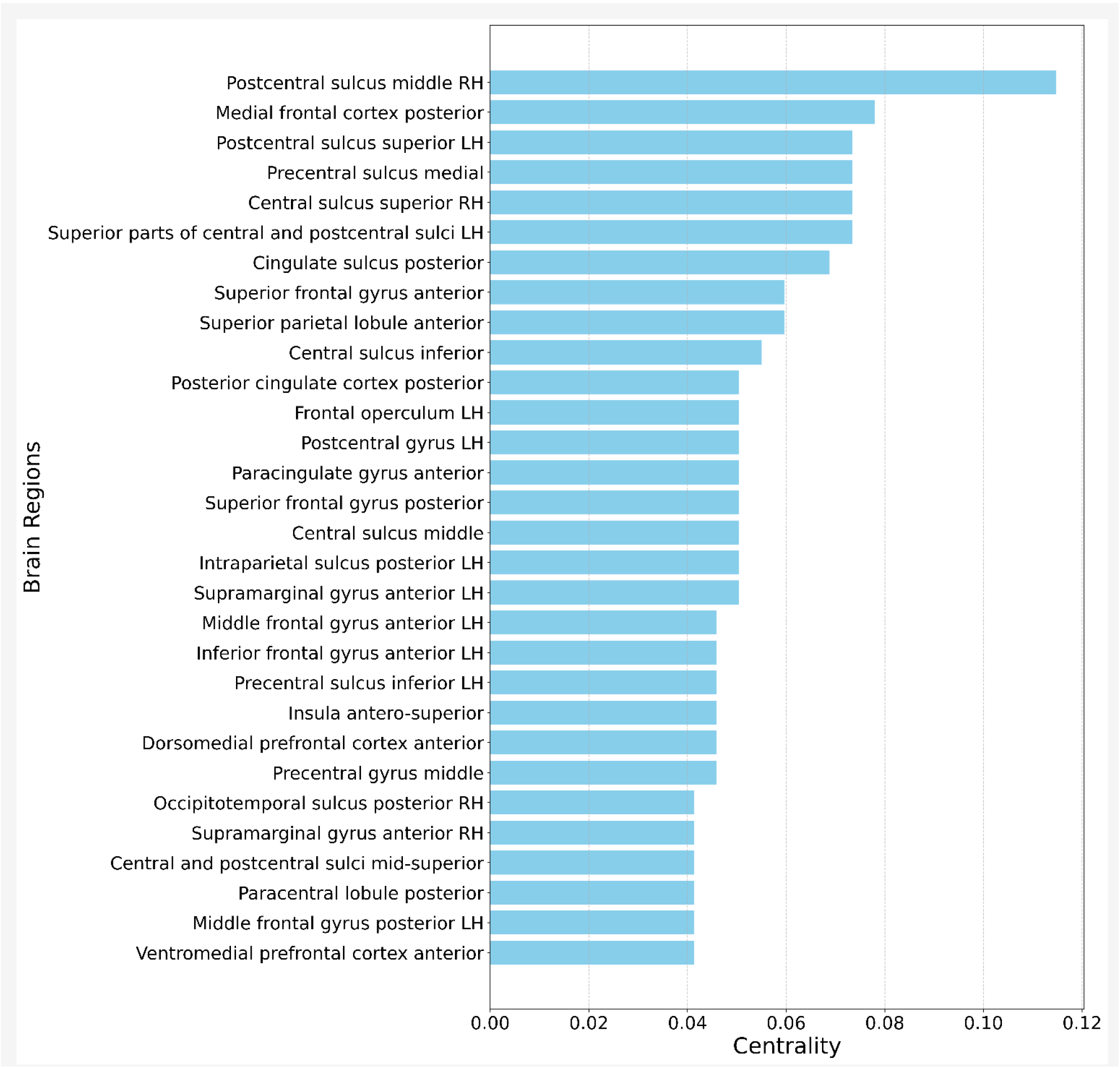
30 highest degree centrality values in identified network (NBS) Top central regions include the insula and parts of prefrontal, cingulate and somatosensory cortices, all implicated in pain literature.

### Anatomical analysis

GLM analysis of whole-brain gray matter volume (FreeSurfer parcellation) in Figure 3 revealed widespread reductions in the pain group compared to pain-free controls, with 44 of 131 brain regions (33.6%) showing posterior distributions entirely below zero, indicating reduced volume in the chronic pain group. In contrast, only 1 region’s interval (0.76%) was entirely above zero. The average normalized gray matter volume difference (pain minus control) was -3.98% ± 1.2% across the 44 regions. Notably, reductions were observed in the ACC, and amygdala. Given their prominent role in the pain literature, the ACC and insula are presented in greater detail in Supplement B, with full reporting of all regressors and sex-related effects. In both regions, gray matter volume was reduced in the pain group across sexes. Although the same analytical approach was applied to all regions, the results for other regions, shown in Figure 3, focus solely on the pain group regressor.

**Figure 3:**
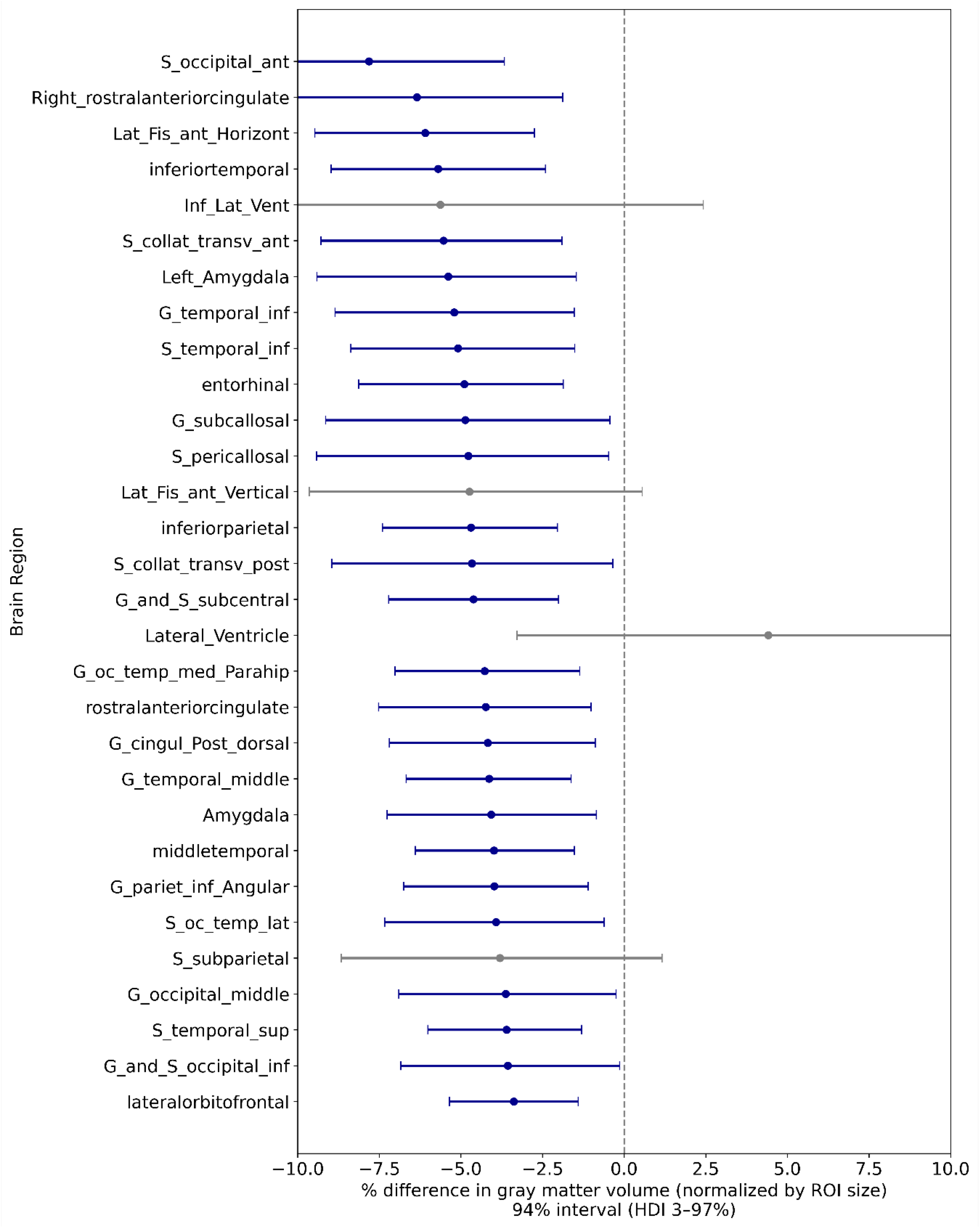
Bayesian anatomical analysis of top 30 regions with largest normalized gray matter volume difference (pain minus control) Whole brain Bayesian analysis showing differences in gray matter volume between groups, normalized by regional size. Each point represents the posterior mean estimate of the group difference for a given region, and the horizontal lines indicate the 94% HDI. Regions are ordered in descending magnitude of percentage difference. Blue intervals indicate regions for which the entire HDI lies below zero, reflecting robust reduced gray matter volume in the pain group relative to controls; gray intervals cross zero. Prominent regions include the cingulate cortex and the amygdala (G: gyrus, S: sulcus).

### Sensitivity analysis: functional analysis

To determine whether the large observed group differences in correlation matrices of functional analysis were driven by noise, two control analyses were performed. The mean absolute correlation within the NBS-identified network was calculated for each participant (excluding the eight noisy participants) and examined in relation to (1) each participant’s mean Derivative of Variance (DVARS) (Figure 4A), where higher values reflect greater instability over time, and (2) the Root Mean Square (RMS, Figure 4B) distance of each participant’s correlation matrix from the group mean, where higher values indicate that an individual’s connectivity profile differs substantially from their group average, and may not represent the group-level pattern.

**Figure 4:**
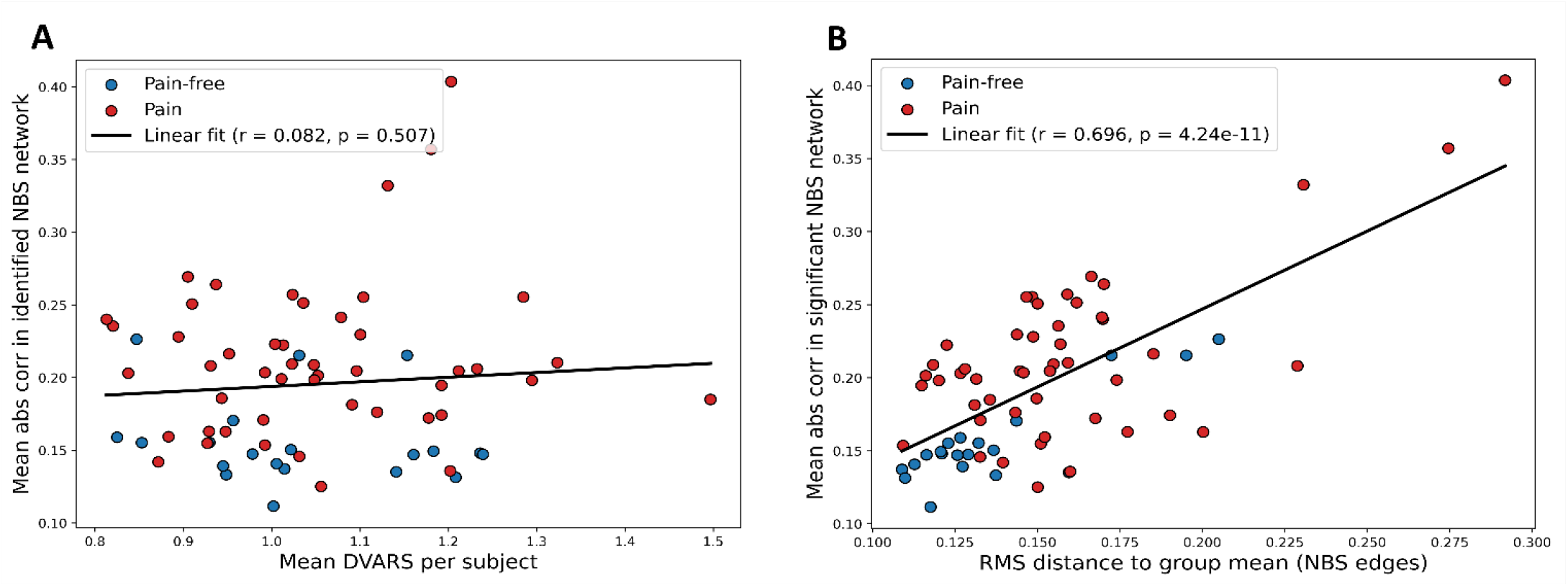
Sensitivity analysis. **(A) Mean abs corr in identifies NBS network vs. mean DVARS** Relationship between data quality and connectivity strength. Mean absolute correlation values showed no meaningful association with DVARS across participants (r=0.082), suggesting that overall connectivity strength was not driven by temporal signal variability. **(B) Mean abs corr in identifies NBS network vs. RMS of single corr matrix and the group’s average** Association between connectivity strength and deviation from the group connectivity pattern. Mean absolute correlation values were strongly correlated with the RMS distance of each participant’s correlation matrix from the group average (r = 0.696), indicating that participants with stronger overall correlations also tended to deviate more from the group-level connectivity structure. After excluding the four participants with the largest RMS distances, the association decreased to a moderate level (r = 0.426).

## DISCUSSION

This study provides further evidence that chronic pain is associated with widespread alterations in both functional connectivity and brain structure. Most notably, *functional* analysis revealed heightened resting-state functional connectivity across widespread brain regions (75% of brain regions) in patients with chronic pain compared to pain-free controls. A major functional network showed strong positive correlations in the pain group and substantially lower or even negative correlations in the pain-free group. Most high-degree centrality nodes contributing to these network-level disturbances include brain regions with established roles in pain processing, interoception, and emotional regulation ^17,19^, including the somatosensory cortex, cingulate cortex, and prefrontal regions. When these regions were mapped onto Yeo’s 7-network parcellation ^48^, one third of the top 30 regions belonged to the somatomotor network, while the remaining regions were distributed across the default mode network (DMN), salience, and dorsal attention networks, highlighting the involvement of multiple higher-order processes beyond primary sensory processing. These findings align with and extend the dynamic pain connectome framework proposed by Kucyi and Davis (2015), which conceptualizes chronic pain as emerging from large-scale neural network dysregulation rather than isolated regional abnormalities. Our whole-brain analysis extends this literature by showing that heightened connectivity is not limited to specific network pairs but represents a more global pattern of enhanced functional connectivity throughout the brain.

Importantly, our findings also revealed widespread *structural* alterations accompanying these functional changes. Structural analyses using a Bayesian whole-brain GLM revealed gray matter reductions in 44 out of 131 cortical and subcortical regions (33.6%), with the largest differences observed in areas including the ACC and amygdala. The ROI-level models showed robust group differences after accounting for age, sex, and total intracranial volume. Given their prominent appearance in the pain literature ^17,49^, detailed results were presented for the insula and ACC (supplement B), both of which showed reduced gray matter volume in the pain group across sexes. These structural changes are consistent with previous reports of gray matter alterations in chronic pain ^15,18^, though the mechanisms underlying these changes, whether they represent neuroplastic adaptation, neuroinflammation, or pre-existing predisposition, remain unclear.

The convergence of functional and structural alterations in overlapping regions suggests coordinated changes across multiple levels of brain organization. Previous longitudinal work has shown progressive connectivity reorganization during the transition from acute to chronic back pain ^19,21^, suggesting that at least some of these changes may develop due to persistent pain rather than representing pre-existing vulnerabilities. However, the cross-sectional nature of our data precludes causal inference, and both interpretations remain plausible. Additional analyses examining pairwise inter-network correlations and predictive models using pain duration and pain intensity as predictors did not reveal reliable or interpretable relationships (see Supplement C).

Since between-subject functional variability was higher in the pain group, we conducted sensitivity analyses to examine whether the connectivity differences could be explained by noise, such as motion or signal fluctuations. Specifically, we examined the relationship between each participant’s mean absolute correlation value in the NBS-identified network and (1) DVARS, reflecting temporal signal instability, and (2) the RMS distance between each participant’s correlation matrix and their group-average matrix. The first analysis showed no meaningful relationship between connectivity strength and DVARS, indicating that temporal signal fluctuations do not explain the group differences. However, the second analysis revealed a strong correlation (Pearson’s r = 0.696) between RMS distance and absolute mean correlation values. Overall, two interpretations for this finding are possible.

The first interpretation is that the RMS-connectivity relationship reflects genuine neural variability: participants whose connectivity patterns deviate more from the group average may do so precisely because their connectivity is stronger, meaning that higher absolute correlations and larger RMS values reflect meaningful individual differences in pain-related network organization rather than artifact. An alternative interpretation is that the observed group differences in connectivity may reflect noise, such that participants who deviate more from the group average, due to motion or other artifacts, also exhibit higher apparent connectivity. However, two observations argue against noise as the primary explanation: (1) DVARS analysis indicated that temporal noise was not the primary contributor, and (2) the correlation dropped from 0.696 to 0.426 when the four most extreme participants were removed, suggesting the effect is not entirely driven by noise. Taken together, these observations are consistent with an interpretation in which the RMS-connectivity relationship reflects structured variability across participants rather than noise, and provides a coherent explanation for the entire distribution of data points, including the four extreme cases. At the same time, alternative explanations related to noise cannot be ruled out, and the present findings should therefore be interpreted cautiously. Future studies with larger samples could examine how functional connectivity in chronic pain relates to deviation from the group mean.

In summary, these findings indicate that chronic pain is characterized by widespread alterations in both functional connectivity and brain structure, extending beyond isolated regions to involve large-scale network organization. By combining whole-brain functional and structural analyses with sensitivity tests addressing noise-related interpretations, this study broadens the network-level conceptualization of chronic pain to include widespread whole-brain alterations. Importantly, identifying consistent patterns of network dysregulation and structural alterations may help refine neurobiological models of chronic pain and support future efforts to identify biomarkers relevant for prognosis, treatment stratification, and monitoring therapeutic change.

### Limitations

Several limitations must be considered when interpreting these results. First, the modest sample size (N=56 pain patients, N=20 controls), while comparable to similar studies, may limit statistical power and generalizability. Second, while rs-fMRI provides valuable information about network-level brain function, it lacks the temporal resolution necessary to capture fast neural dynamics, and its interpretation remains correlational. Third, chronic pain patients often present with comorbid depression or anxiety (prevalence is estimated at about 40% of chronic pain patients ^50^), which could independently affect brain connectivity patterns. Fourth, since MRI scans were obtained only before the intervention, we cannot evaluate how each person’s brain activity changed over time, as no baseline comparison is available. Lastly, because this is a cross-sectional study, we cannot determine whether chronic pain causes the observed structural and functional differences or whether individuals with particular neural architectures are more susceptible to developing persistent pain. Longitudinal studies tracking patients from acute injury through the development of chronic pain are essential to establish the temporal sequence of these changes and identify potential predictive biomarkers.

## CONCLUSIONS

Taken together, these findings demonstrate that chronic pain is associated with broad and distributed alterations across the brain involving both connectivity dynamics and gray-matter structure, particularly in regions supporting interoceptive, affective, and cognitive aspects. The heightened functional connectivity observed in pain patients, localized to regions consistently implicated in pain literature across independent studies, extends the dynamic pain connectome framework and provides new evidence for network-level dysregulation as a core feature of chronic pain. The convergence of functional and structural changes in overlapping pain-processing regions strengthens the case for chronic pain as a disorder involving coordinated alterations across multiple levels of brain organization. These results reinforce emerging network-based perspectives of chronic pain, and highlight the promise of resting-state fMRI as a non-invasive and accessible tool for identifying objective neural markers; an important step toward improving diagnosis, prognosis, and treatment monitoring.

To translate these findings into clinical utility, future research must employ larger, potentially multisite, and longitudinal samples to replicate these findings, establish their specificity to chronic pain versus other conditions, and determine whether the observed brain changes precede or arise from chronic pain. If validated, these neural signatures could reliably predict individual treatment outcomes and support the development of personalized approaches to pain management, ultimately addressing a major unmet clinical need.

## Data Availability

The minimum dataset necessary to interpret and verify the findings of this study is publicly available on OpenNeuro at doi:10.18112/openneuro.ds000208.v1.0.1. This includes the neuroimaging data used in the analyses. The analysis code is openly available at
www.gitlab.com/dueklab/10_programs/chronic-pain-mri-analysis-openneuro
All derived data and demographic data are available from the corresponding author upon request and approval of the investigator who collected the data.

https://gitlab.com/dueklab/10_programs/chronic-pain-mri-analysis-openneuro

## Data availability

The minimum dataset necessary to interpret and verify the findings of this study is publicly available on OpenNeuro at doi:10.18112/openneuro.ds000208.v1.0.1. This includes the neuroimaging data used in the analyses. The analysis code is openly available at www.gitlab.com/dueklab/10_programs/chronic-pain-mri-analysis-openneuro/

All derived data and demographic data are available from the corresponding author upon request and approval of the investigator who collected the data.

## REFERENCES

1. Kumbhare, D. A., Elzibak, A. H. & Noseworthy, M. D. Evaluation of Chronic Pain Using Magnetic Resonance (MR) Neuroimaging Approaches: What the Clinician Needs to Know. Clin. J. Pain 33, 281–290 (2017).

2. Varrassi, G. et al. Pharmacological treatment of chronic pain – the need for CHANGE. Curr. Med. Res. Opin. 26, 1231–1245 (2010).

3. Chenaf, C. et al. Prevalence of chronic pain with or without neuropathic characteristics in France using the capture-recapture method: a population-based study: a population-based study. Pain 159, 2394–2402 (2018).

4. Santiago, B. V. M. et al. Prevalence of chronic pain in Brazil: A systematic review and meta-analysis. Clinics (Sao Paulo) 78, 100209 (2023).

5. Yong, R. J., Mullins, P. M. & Bhattacharyya, N. Prevalence of chronic pain among adults in the United States. Pain 163, e328–e332 (2022).

6. Fayaz, A., Croft, P., Langford, R. M., Donaldson, L. J. & Jones, G. T. Prevalence of chronic pain in the UK: a systematic review and meta-analysis of population studies. BMJ Open 6, e010364 (2016).

7. Sharon, H., Greener, H., Hochberg, U. & Brill, S. The Prevalence of Chronic Pain in the Adult Population in Israel: An Internet-Based Survey. Pain Res. Manag. 2022, 3903720 (2022).

8. Wager, T. D. Managing Pain. Cerebrum 2022, (2022).

9. Zhang, Z., Gewandter, J. S. & Geha, P. Brain imaging biomarkers for chronic pain. Front. Neurol. 12, 734821 (2021).

10. Basbaum, A. I., Jensen, T. S. & Keefe, F. J. Fifty years of pain research and clinical advances: highlights and key trends. Pain 164, S11–S15 (2023).

11. Melzack, R. From the gate to the neuromatrix. Pain 82, S121–S126 (1999).

12. Loeser, J. D. & Melzack, R. Pain: an overview. Lancet 353, 1607–1609 (1999).

13. Kucyi, A. & Davis, K. D. The dynamic pain connectome. Trends Neurosci. 38, 86–95 (2015).

14. Mills, E. P. et al. Sex-Specific White Matter Abnormalities Across the Dynamic Pain Connectome in Neuropathic Pain: A Fixel-Based Analysis Study. Human Brain Mapping 46, e70135 (2025).

15. Wager, T. D., Zorina-Lichtenwalter, K. & Friedman, N. P. A New Look at Gray Matter Decreases in Chronic Pain. Biol. Psychiatry 95, 387–388 (2024).

16. Farmer, M. A., Baliki, M. N. & Apkarian, A. V. A dynamic network perspective of chronic pain. Neurosci. Lett. 520, 197– 203 (2012).

17. Jensen, K. B. et al. Brain activations during pain: a neuroimaging meta-analysis of patients with pain and healthy controls. Pain 157, 1279–1286 (2016).

18. Geha, P. Y. et al. The brain in chronic CRPS pain: abnormal gray-white matter interactions in emotional and autonomic regions. Neuron 60, 570–581 (2008).

19. Baliki, M. N. et al. Corticostriatal functional connectivity predicts transition to chronic back pain. Nat. Neurosci. 15, 1117–1119 (2012).

20. Apkarian, V. A., Hashmi, J. A. & Baliki, M. N. Pain and the brain: specificity and plasticity of the brain in clinical chronic pain. Pain 152, S49–S64 (2011).

21. Hashmi, J. A. et al. Shape shifting pain: chronification of back pain shifts brain representation from nociceptive to emotional circuits. Brain 136, 2751–2768 (2013).

22. Khosla, M., Jamison, K., Ngo, G. H., Kuceyeski, A. & Sabuncu, M. R. Machine learning in resting-state fMRI analysis. Magn. Reson. Imaging 64, 101–121 (2019).

23. Parkes, L., Satterthwaite, T. D. & Bassett, D. S. Towards precise resting-state fMRI biomarkers in psychiatry: synthesizing developments in transdiagnostic research, dimensional models of psychopathology, and normative neurodevelopment. Curr. Opin. Neurobiol. 65, 120–128 (2020).

24. Filippi, M. et al. The human functional connectome in neurodegenerative diseases: relationship to pathology and clinical progression. Expert Rev. Neurother. 23, 59–73 (2023).

25. Finn, E. S. Is it time to put rest to rest? Trends Cogn. Sci. 25, 1021–1032 (2021).

26. Lurie, D. J. et al. Questions and controversies in the study of time-varying functional connectivity in resting fMRI. Netw. Neurosci. 4, 30–69 (2020).

27. Santana, A. N., Cifre, I., de Santana, C. N. & Montoya, P. Using deep learning and resting-state fMRI to classify chronic pain conditions. Front. Neurosci. 13, 1313 (2019).

28. Tétreault, P. et al. Brain Connectivity Predicts Placebo Response across Chronic Pain Clinical Trials. PLoS Biol. 14, e1002570 (2016).

29. Tétreault, P. et al. Inferring distinct mechanisms in the absence of subjective differences: Placebo and centrally acting analgesic underlie unique brain adaptations. Hum. Brain Mapp. 39, 2210–2223 (2018).

30. Esteban, O. et al. fMRIPrep: a robust preprocessing pipeline for functional MRI. Nat. Methods 16, 111–116 (2019).

31. Yang, J., Gohel, S. & Vachha, B. Current methods and new directions in resting state fMRI. Clin. Imaging 65, 47–53 (2020).

32. Dadi, K. et al. Fine-grain atlases of functional modes for fMRI analysis. Neuroimage 221, 117126 (2020).

33. Bullmore, E. & Sporns, O. Erratum: Complex brain networks: graph theoretical analysis of structural and functional systems. Nat. Rev. Neurosci. 10, 312–312 (2009).

34. Thompson, W. H. & Fransson, P. On stabilizing the variance of dynamic functional brain connectivity time series. Brain Connect. 6, 735–746 (2016).

35. Zalesky, A., Fornito, A. & Bullmore, E. T. Network-based statistic: identifying differences in brain networks. Neuroimage 53, 1197–1207 (2010).

36. Fischl, B. FreeSurfer. Neuroimage 62, 774–781 (2012).

37. Dale, A. M., Fischl, B. & Sereno, M. I. Cortical surface-based analysis. I. Segmentation and surface reconstruction. Neuroimage 9, 179–194 (1999).

38. Keller, S. S. et al. Volume estimation of the thalamus using freesurfer and stereology: consistency between methods. Neuroinformatics 10, 341–350 (2012).

39. Sánchez-Benavides, G. et al. Manual validation of FreeSurfer’s automated hippocampal segmentation in normal aging, mild cognitive impairment, and Alzheimer Disease subjects. Psychiatry Res. 181, 219–225 (2010).

40. Cardinale, F. et al. Validation of FreeSurfer-estimated brain cortical thickness: comparison with histologic measurements. Neuroinformatics 12, 535–542 (2014).

41. Ávila, H., Raulino Silva, V. & Magalhães, D. S. F. Volume estimation of the brain, white matter, and gray matter using FreeSurfer and FSL: consistency between methods. Res. Biomed. Eng. 35, 257–263 (2019).

42. Coventry, B. S. & Bartlett, E. L. Practical Bayesian inference in neuroscience: Or how I learned to stop worrying and embrace the distribution. eNeuro 11, ENEURO.0484–23.2024 (2024).

43. Korem, N. et al. Post-treatment alterations in white matter integrity in PTSD: Effects on symptoms and functional connectivity a secondary analysis of an RCT. Psychiatry Res. Neuroimaging 343, 111864 (2024).

44. Kruschke, J. K. Rejecting or accepting parameter values in Bayesian estimation. Adv. Methods Pract. Psychol. Sci. 1, p270–280 (2018).

45. Mcelreath, R. rethinking: Statistical Rethinking book package. R package version 1, p(2014).

46. Kumar, R., Carroll, C., Hartikainen, A. & Martin, O. ArviZ a unified library for exploratory analysis of Bayesian models in Python. J. Open Source Softw. 4, 1143 (2019).

47. McElreath, R. Statistical Rethinking. (CRC Press, London, England, 2020). doi:10.1201/9780429029608.

48. Yeo, B. T. T. et al. The organization of the human cerebral cortex estimated by intrinsic functional connectivity. J. Neurophysiol. 106, 1125–1165 (2011).

49. Zeng, X., Sun, Y., Zhiying, Z., Hua, L. & Yuan, Z. Chronic pain-induced functional and structural alterations in the brain: A multi-modal meta-analysis. J. Pain 28, 104740 (2025).

50. Aaron, R. V. et al. Prevalence of depression and anxiety among adults with chronic pain: A systematic review and meta-analysis: A systematic review and meta-analysis. JAMA Netw. Open 8, e250268 (2025).

