## Supplemental data for "Neural Alterations in Chronic Pain: MRI Analysis"

### SUPPLEMENTS

#### Supplement A

| Variable | Study 1 (Placebo) Responder | Study 1 (Placebo) Non-Responder | p-value | Study 2 (Placebo) Responder | Study 2 (Placebo) Non-Responder | p-value | Study 2 (Duloxetine) Responder | Study 2 (Duloxetine) Non-Responder | p-value | Controls |
|---|---|---|---|---|---|---|---|---|---|---|
| Sex (F/M) | 5F/3M | 4F/5M | — | 7F/4M | 5F/5M | — | 5F/3M | 5F/6M | — | 10F/10M |
| Age (years) | 55.9 (1.5) | 57.8 (2.3) | 0.51 | 54.7 (3.0) | 62.0 (2.6) | 0.08 | 57.6 (0.8) | 60.3 (1.7) | 0.17 | 57.9 (1.5) |
| Pain Duration (years) | 11.4 (3.6) | 13.3 (3.8) | 0.72 | 10.2 (2.8) | 12.7 (3.4) | 0.57 | 7.3 (2.4) | 11.6 (3.1) | 0.23 | — |
| BDI | 3.4 (1.5) | 5.2 (1.4) | 0.39 | 2.7 (1.2) | 10.5 (3.6) | 0.07 | 3.8 (1.6) | 7.6 (3.3) | 0.17 | 2.5 (2.2) |
| PCS | 25.0 (5.4) | 16.9 (4.5) | 0.27 | 9.0 (2.2) | 24.5 (4.2) | 0.006 | 5.4 (0.9) | 18.3 (4.8) | 0.03 | — |
| MQS | 7.1 (0.7) | 7.0 (0.6) | 0.91 | 4.7 (2.2) | 10.0 (3.8) | 0.25 | 5.1 (2.5) | 10.6 (4.8) | 0.32 | — |

Table 1: S1 table supplement, demographics and pain scores for participants in study 1, study 2, and controls [28].

#### Supplement B: anatomical analysis- insula and ACC gray matter volume

Insula (Figure S1) and ACC (Figure S2) show substantial gray matter volume reductions in the pain group, across sexes.

**Anatomical analysis: Insula gray matter volume**

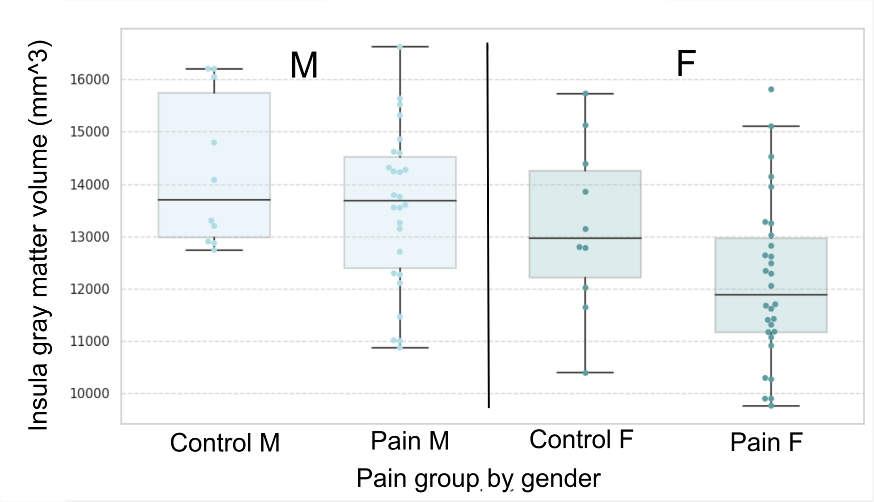


Figure S1- GLM analysis- mean gray matter volume difference, Bayesian approach (94% credible interval): **Pain group: -331mm$^3$** (-619, -35), **Total intracranial volume: 1009mm$^3$** (600,1415), Gender: -3mm$^3$ (-412, 398), Age: -171mm$^3$ (-482, 124)

**Anatomical analysis: rostral ACC gray matter volume**

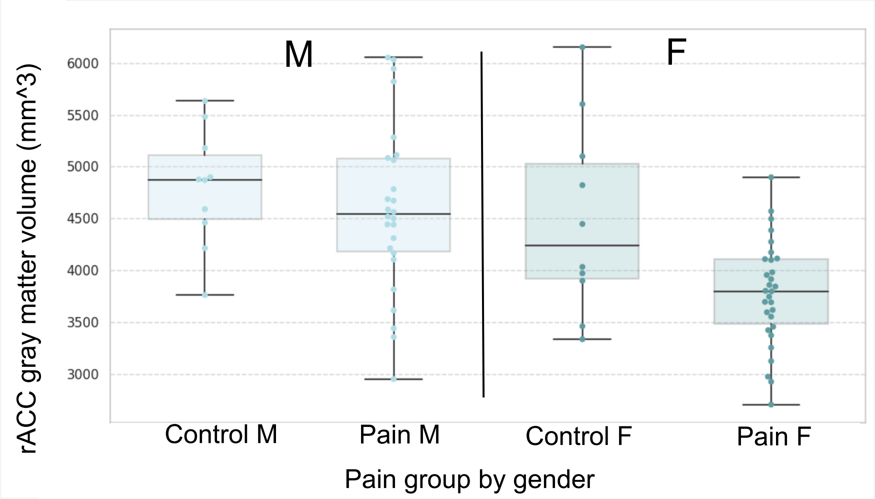


Figure S2- GLM analysis- mean gray matter volume difference, Bayesian approach (94% credible interval): **Pain group: -180mm$^3$** (97% CrI: -326, -44), **Total intracranial volume: 391mm$^3$** (CrI: 195,581), Age: -15mm$^3$ (CrI: -165, 123), Gender: 83mm$^3$ (CrI: -107, 272)

#### Supplement C: additional analyses

Additional functional analyses not included in the manuscript examined potential brain-behavior associations; no reliable predictive relationships were detected. First, to examine the correlation between gray matter volume deterioration and pain duration (in years), a linear regression analysis was employed, with regressors for age, sex, and total intracranial volume, focusing on the top 5 regions showing the greatest gray matter volume difference between groups. Second, Connectome-based Predictive Modeling (CPM) and machine-learning regression (XGBoost) were applied to predict pain levels from individual functional connectivity patterns. Third, network-level correlations based on Yeo's 7-network parcellation did not reveal significant or interpretable patterns when examining pairwise correlations between the networks. These findings suggest that although chronic pain is associated with broad alterations in connectivity, the relationship between network architecture and pain intensity may be individual, non-linear, context-dependent, or influenced by factors not captured in resting-state data. For details, refer to our GitHub repository:
www.gitlab.com/dueklab/10_programs/chronic-pain-mri-analysis-openneuro/

#### Supplement D: fmriprep documentation

Results included in this manuscript come from preprocessing performed using *fMRIPrep* 24.0.1 (@fmriprep1; @fmriprep2; RRID:SCR_016216), which is based on *Nipype* 1.8.6

(@nipype1; @nipype2; RRID:SCR_002502).

**Anatomical data preprocessing**

A total of 1 T1-weighted (T1w) images were found within the input :

BIDS dataset. The T1w image was corrected for intensity non-uniformity (INU) with `N4BiasFieldCorrection` [@n4], distributed with ANTs 2.5.1 [@ants, RRID:SCR_004757], and used as T1w-reference throughout the workflow.

The T1w-reference was then skull-stripped with a *Nipype* implementation of the `antsBrainExtraction.sh` workflow (from ANTs), using OASIS30ANTs as target template.

Brain tissue segmentation of cerebrospinal fluid (CSF), white-matter (WM) and gray-matter (GM) was performed on the brain-extracted T1w using `fast` [FSL (version unknown), RRID:SCR_002823, @fsl_fast].

Brain surfaces were reconstructed using `recon-all` [FreeSurfer 7.3.2, RRID:SCR_001847, @fs_reconall], and the brain mask estimated previously was refined with a custom variation of the method to reconcile ANTs-derived and FreeSurfer-derived segmentations of the cortical gray-matter of Mindboggle [RRID:SCR_002438, @mindboggle].

Volume-based spatial normalization to one standard space (MNI152NLin2009cAsym) was performed through nonlinear registration with `antsRegistration` (ANTs 2.5.1), using brain-extracted versions of both T1w reference and the T1w template.

The following template was were selected for spatial normalization and accessed with *TemplateFlow* [24.2.0, @templateflow]:

*ICBM 152 Nonlinear Asymmetrical template version 2009c* [@mni152nlin2009casym, RRID:SCR_008796; TemplateFlow ID: MNI152NLin2009cAsym].

**Functional data preprocessing**

For each of the 1 BOLD runs found per subject (across all tasks and sessions), the following preprocessing was performed.

First, a reference volume was generated, using a custom methodology of *fMRIPrep*, for use in head motion correction.

Head-motion parameters with respect to the BOLD reference (transformation matrices, and six corresponding rotation and translation parameters) are estimated before any spatiotemporal filtering using `mcflirt` [FSL <ver>, @mcflirt].

The BOLD reference was then co-registered to the T1w reference using `bbregister` (FreeSurfer) which implements boundary-based registration [@bbr].

Co-registration was configured with six degrees of freedom.

Several confounding time-series were calculated based on the *preprocessed BOLD*: framewise displacement (FD), DVARS and three region-wise global signals.

FD was computed using two formulations following Power (absolute sum of relative motions, @power_fd_dvars) and Jenkinson (relative root mean square displacement between affines, @mcflirt).

FD and DVARS are calculated for each functional run, both using their implementations in *Nipype* [following the definitions by @power_fd_dvars].

The three global signals are extracted within the CSF, the WM, and the whole-brain masks.

Additionally, a set of physiological regressors were extracted to allow for component-based noise correction [*CompCor*, @compcor].

Principal components are estimated after high-pass filtering the *preprocessed BOLD* time-series (using a discrete cosine filter with 128s cut-off) for the two *CompCor* variants: temporal (tCompCor) and anatomical (aCompCor).

tCompCor components are then calculated from the top 2% variable voxels within the brain mask.

For aCompCor, three probabilistic masks (CSF, WM and combined CSF+WM) are generated in anatomical space.

The implementation differs from that of Behzadi et al. in that instead of eroding the masks by 2 pixels on BOLD space, a mask of pixels that likely contain a volume fraction of GM is subtracted from the aCompCor masks.

This mask is obtained by dilating a GM mask extracted from the FreeSurfer's *aseg* segmentation, and it ensures components are not extracted from voxels containing a minimal fraction of GM.

Finally, these masks are resampled into BOLD space and binarized by thresholding at 0.99 (as in the original implementation).

Components are also calculated separately within the WM and CSF masks. For each CompCor decomposition, the *k* components with the largest singular values are retained, such that the retained components' time series are sufficient to explain 50 percent of variance across the nuisance mask (CSF, WM, combined, or temporal). The remaining components are dropped from consideration.

The head-motion estimates calculated in the correction step were also placed within the corresponding confounds file.

The confound time series derived from head motion estimates and global signals were expanded with the inclusion of temporal derivatives and quadratic terms for each [@confounds_satterthwaite_2013].

Frames that exceeded a threshold of 0.5 mm FD or 1.5 standardized DVARS were annotated as motion outliers.

Additional nuisance timeseries are calculated by means of principal components analysis of the signal found within a thin band (*crown*) of voxels around the edge of the brain, as proposed by [@patriat_improved_2017].

All resamplings can be performed with *a single interpolation step* by composing all the pertinent transformations (i.e. head-motion transform matrices, susceptibility distortion correction when available, and co-registrations to anatomical and output spaces).

Gridded (volumetric) resamplings were performed using `nitransforms`, configured with cubic B-spline interpolation.

Many internal operations of *fMRIPrep* use *Nilearn* 0.10.4 [@nilearn, RRID:SCR_001362],

mostly within the functional processing workflow. For more details of the pipeline, see [the section corresponding to workflows in *fMRIPrep*'s documentation]
(https://fmriprep.readthedocs.io/en/latest/workflows.html "FMRIPrep's documentation").

##### Copyright Waiver

The above boilerplate text was automatically generated by fMRIPrep with the express intention that users should copy and paste this text into their manuscripts *unchanged*.

It is released under the [CC0](https://creativecommons.org/publicdomain/zero/1.0/) license.
